# Persistent Hypertension After Asymptomatic Elevated In-Hospital Blood Pressure: The INDEBP Study

**DOI:** 10.64898/2026.10.08.26365097

**Authors:** Katharina Hausknecht, Michael Mayr, Tobias Breidthardt, Thilo Burkard, Annina S. Vischer

**Author notes:** Correspondence to PD Dr. med. Annina S. Vischer, Hypertension Centre, ESH Hypertension Centre of Excellence, Medical Outpatient Department, University Hospital Basel, 4031 Basel, Switzerland.

## Abstract

**Background:** Elevated blood pressure (BP) is common during hospitalization. Despite recommendations to avoid routine changes in antihypertensive therapy, clinical practice remains variable. Prospective data on whether elevated in-hospital BP identifies patients with hypertension after discharge are limited.

**Methods:** Adults hospitalized for non-cardiac conditions with asymptomatic routine in-hospital BP ≥140 and/or ≥90 mmHg on at least two occasions separated by ≥24 hours were prospectively enrolled. Antihypertensive therapy was managed at the discretion of treating physicians. Routine in-hospital BP, standardized research BP, and changes in antihypertensive therapy were recorded. Four weeks after discharge, participants underwent 24-hour ambulatory BP monitoring (ABPM).

**Results:** Among 50 participants, 42 (84%; 95% CI, 70.9 – 92.8) had hypertension on follow-up ABPM. Among 21 participants without known hypertension, 18 (86%) had previously unrecognized hypertension on follow-up ABPM; among 29 participants with known hypertension, 24 (83%) had uncontrolled hypertension. Antihypertensive therapy was changed during hospitalization in 26 participants (52%), with de-escalation more frequent than intensification. Hypertension on follow-up ABPM was present in 88.5% of participants with and 79.2% without in-hospital therapy changes. Standardized research BP, but not routine in-hospital BP, correlated with follow-up ABPM.

**Conclusion:** Asymptomatic elevated routine in-hospital BP identified a population with a high prevalence of hypertension four weeks after discharge, including patients with and without previously diagnosed hypertension. These findings support structured post-discharge BP reassessment rather than assuming that elevated in-hospital BP is transient.

URL: www.ClinicalTrials.gov; Unique identifier: NCT06132451

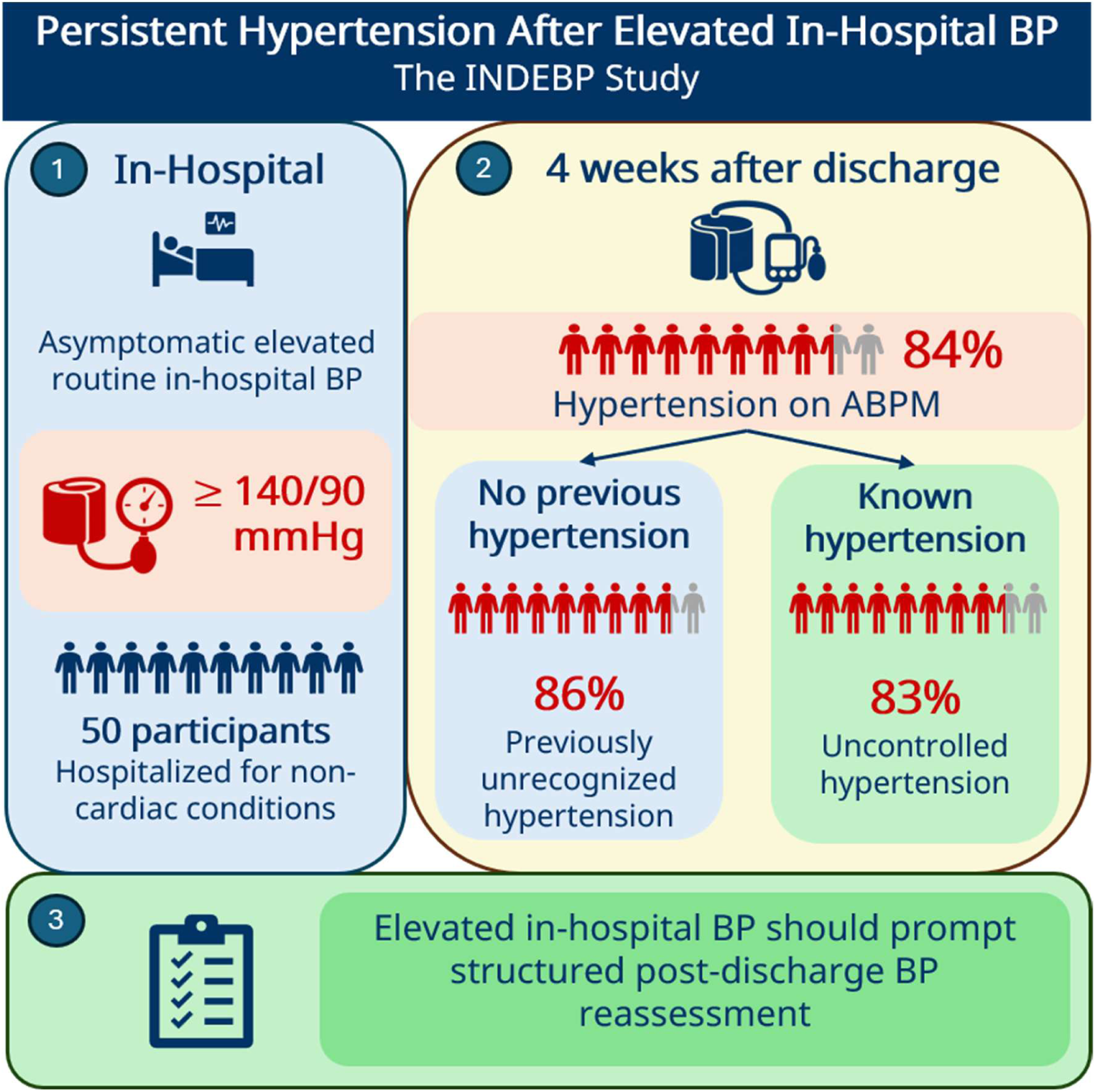

## 1. Introduction

Elevated routine blood pressure (BP) measurements are common during hospitalization, affecting up to 70% of medical inpatients^1, 2^. Although often attributed to acute illness, pain, anxiety, or other transient physiological stressors, it remains uncertain whether asymptomatic elevated in-hospital BP primarily reflects temporary BP elevations or previously unrecognized or uncontrolled hypertension requiring long-term management^2–8^. At the same time, hypertension remains substantially underdiagnosed and undertreated worldwide, with considerable delays between disease onset and diagnosis, partly reflecting clinical inertia and missed opportunities for case identification^9–12^. Hospitalization therefore represents an important opportunity to identify patients with previously unrecognized or uncontrolled hypertension - an opportunity that may currently be underutilized.

This uncertainty has important implications for clinical practice. In the absence of hypertensive emergencies, physicians must balance elevated BP against competing priorities related to the acute illness, including impaired kidney function, infection, volume depletion, and polypharmacy^6, 13^. Consequently, management of asymptomatic elevated in-hospital BP remains highly variable^2, 7^. While some clinicians initiate or intensify antihypertensive therapy, others deliberately avoid treatment changes because elevated BP is considered a transient phenomenon. Recent recommendations, including the American Heart Association Scientific Statement and the Swiss *smarter medicine* initiative, appropriately advise against routine changes in antihypertensive therapy during hospitalization in the absence of hypertensive emergencies^6, 14^. However, these recommendations were necessarily developed in the context of limited prospective evidence regarding the clinical significance of elevated routine in-hospital BP.

Recently, the SHINE study demonstrated that elevated in-hospital BP frequently identifies patients with previously unrecognized hypertension after discharge, supporting the concept that elevated in-hospital BP should not simply be dismissed as a transient phenomenon^15^. However, SHINE primarily evaluated the diagnostic accuracy of in-hospital BP measurements among previously untreated individuals and did not address the broader clinical implications of elevated in-hospital BP in routine clinical practice, including physician management, treatment changes during hospitalization, or the persistence of hypertension in patients with and without previously diagnosed hypertension.

Several clinically relevant questions therefore remain unanswered. Does asymptomatic elevated routine in-hospital BP predict previously unrecognized or uncontrolled hypertension after discharge? Does standardized BP measurement provide additional information regarding subsequent ambulatory BP^16, 17^? How are these patients currently managed during hospitalization? Are changes in antihypertensive therapy associated with BP status after discharge? Addressing these questions is essential for developing evidence-based diagnostic and therapeutic pathways for patients with asymptomatic elevated in-hospital BP.

The prospective In-Hospital Detection of Elevated Blood Pressure (INDEBP) study was designed to evaluate the clinical significance of asymptomatic elevated routine in-hospital BP^18^. In this analysis of the INDEBP study, we aimed primarily to determine the prevalence of previously unrecognized or uncontrolled hypertension on standardized ABPM four weeks after hospital discharge among patients with repeated asymptomatic elevated routine in-hospital BP. Secondary aims were to characterize changes in antihypertensive therapy during hospitalization and their association with follow-up BP status, and to compare routine and standardized in-hospital BP measurements and their associations with subsequent ambulatory BP.

## 2. Methods

### 2.1. Study design and participants

The In-Hospital Detection of Elevated Blood Pressure (INDEBP) study is a prospective, open-label, single-center study conducted at the University Hospital Basel, Switzerland. The detailed protocol has been published previously^18^. The study protocol allowed for an interim assessment of study assumptions during recruitment to evaluate feasibility and the need for potential sample size adjustment. An interim analysis was conducted after 50 participants had completed follow-up including ABPM. In view of the unexpectedly high prevalence of hypertension observed at this assessment and the challenges encountered with recruitment and follow-up, further recruitment to this study phase was discontinued. The present report therefore represents the final analysis of this study phase.

The study was approved by the Ethics Committee for northwest and central Switzerland (EKNZ 2023-01566) and registered on www.ClinicalTrials.gov (NCT06132451). All participants provided written informed consent.

Adult patients hospitalized for non-cardiac conditions were eligible if routine in-hospital BP was systolic BP ≥140 mmHg and/or diastolic BP ≥90 mmHg on at least two occasions separated by ≥24 hours. The study team was automatically notified by the clinical information system when patients hospitalized on the internal medicine wards met these BP criteria. Patients identified through these alerts were subsequently screened for eligibility by the study team. Patients with symptomatic or severe hypertension (systolic BP ≥180 mmHg and/or diastolic BP ≥110 mmHg), hospitalization for acute cardiovascular conditions (stroke, acute coronary syndrome, or decompensated heart failure), pregnancy, age <18 years, inability to undergo ABPM, or inability to provide informed consent were excluded.

In this study, the term “elevated blood pressure” refers to routine in-hospital systolic BP ≥140 mmHg and/or diastolic BP ≥90 mmHg measured during hospitalization and should not be confused with the ESC 2024 BP category “elevated BP”^19^.

### 2.2. Study procedures

#### 2.2.1. Blood pressure assessment

Routine in-hospital BP measurements were performed by ward nurses according to local clinical practice using validated oscillometric devices. For study purposes, the mean routine in-hospital BP was calculated as an average of the two elevated BP measurements that resulted in inclusion.

Standardized research BP was measured by trained study personnel during the baseline visit using a validated automated oscillometric device (WatchBP Office AFIB, Microlife AG, Widnau, Switzerland). After 5 minutes of seated rest, three consecutive BP measurements were obtained at 1-minute intervals. The mean of the second and third measurements was used for analysis. For both routine in-hospital and standardized research BP, elevated BP was defined as a mean systolic BP ≥140 mmHg and/or diastolic BP ≥90 mmHg. For simplicity, this will be expressed as BP ≥140/90 mmHg throughout the manuscript.

All participants underwent 24-hour ABPM four weeks after hospital discharge using validated oscillometric monitors (Mobil-O-Graph NG, I.E.M., Stolberg, Germany). BP measurements were obtained every 20 minutes during daytime and every 30 minutes during nighttime. Awake and asleep periods were defined according to individual patient diaries. Hypertension was defined according to current ESC recommendations as a mean 24-hour systolic BP ≥130 and/or diastolic BP ≥80 mmHg, awake systolic BP ≥135 and/or diastolic BP ≥85 mmHg, or asleep systolic BP ≥120 and/or diastolic BP ≥70 mmHg^19^.

#### 2.2.2. Antihypertensive therapy and follow-up

Changes in antihypertensive therapy during hospitalization were documented prospectively and included treatment initiation, discontinuation, dose escalation, or dose reduction between hospital admission and discharge.

At the follow-up visit four weeks after discharge, ABPM results, antihypertensive therapy, adverse events, and rehospitalizations were assessed. Changes in antihypertensive therapy initiated by primary care physicians after hospital discharge were recorded separately.

#### 2.2.3. Clinical data collection

Baseline demographic characteristics, medical history, medication, laboratory values, and available imaging findings were obtained from standardized patient interviews and electronic medical records. Comorbidities were recorded according to the diagnoses documented in the electronic medical record and were not additionally adjudicated based on laboratory, imaging, or other clinical findings. Pain and anxiety during hospitalization were assessed using routine numerical rating scales (0–10).

### 2.3. Outcomes

The primary endpoint was the prevalence of hypertension on follow-up ABPM. Previously unrecognized hypertension was defined as hypertension on follow-up ABPM in participants without a previous diagnosis of hypertension, whereas uncontrolled hypertension was defined as hypertension on follow-up ABPM in participants with previously diagnosed hypertension.

Secondary endpoints included (1) changes in antihypertensive therapy during hospitalization, (2) comparison of characteristics between participants with and without changes in antihypertensive therapy, (3) comparison of routine in-hospital and standardized research BP measurements, (4) associations of routine in-hospital and standardized research BP with follow-up ABPM.

Prespecified safety endpoints comprised hypertensive complications (intracerebral hemorrhage, ischemic stroke, myocardial infarction, cardiovascular death, and cardiovascular rehospitalization) and hypotensive complications (falls, acute kidney injury, electrolyte disturbances, symptomatic hypotension, and hypotension-related rehospitalization). During hospitalization, hypotension was defined as more than three systolic BP measurements <90 mmHg, consistent with previous definitions^20^.

### 2.4. Statistical analysis

Continuous variables are presented as median (interquartile range [IQR]) and categorical variables as number (%). Between-group comparisons were performed using the Mann-Whitney U test for continuous variables and the χ² test or Fisher exact test for categorical variables, as appropriate. Routine in-hospital and standardized research BP measurements were compared within participants using paired t tests, separately for systolic and diastolic BP. Mean paired differences are reported with 95% confidence intervals (CIs). Associations of routine in-hospital and standardized research BP with corresponding 24-hour ambulatory BP were assessed using Pearson correlation coefficients with 95% CIs after visual inspection of scatterplots for approximate linearity and influential observations. Spearman rank correlations were performed as sensitivity analyses. In exploratory analyses, routine in-hospital and standardized research BP were entered simultaneously into separate linear regression models for systolic and diastolic 24-hour ambulatory BP. Model assumptions and influential observations were assessed by inspection of diagnostic plots. Unstandardized regression coefficients (B) are reported with 95% CIs. All statistical tests were two-sided, and a *P* value <0.05 was considered statistically significant. Statistical analyses were performed using IBM SPSS Statistics, version 28.0.1.0. (IBM Corp., Armonk, NY, USA), and R, version 4.5.2 (R Foundation for Statistical Computing, Vienna, Austria).

## 3. Results

### 3.1. Study population

Between November 16^th^, 2023 and July 16^th^, 2024, 472 hospitalized patients were screened, of whom 89 met the inclusion criteria and were enrolled. Of these, 50 participants had completed follow-up including post-discharge ABPM (Figure 1).

**Figure 1.**
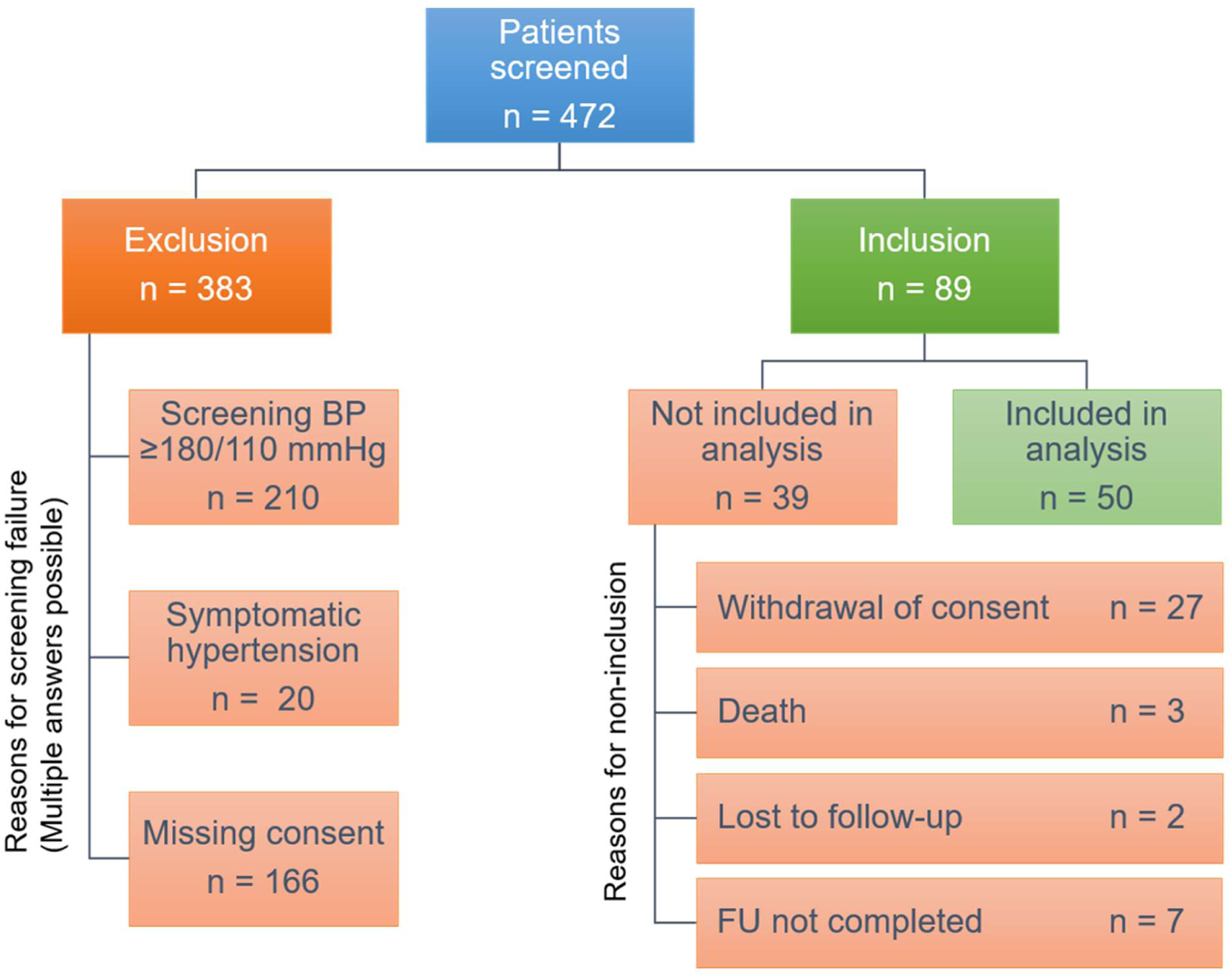
Study flow diagram. Flow diagram showing participant screening, enrollment, exclusions, follow-up, and inclusion in the present analysis. Multiple reasons for exclusion were possible. ABPM = ambulatory blood pressure monitoring; BP = blood pressure; FU = follow-up.

Baseline characteristics are summarized in Table 1 and supplemental Table S1. Participants had a median age of 69.5 years (IQR 62.0 – 77.0), 19 (38%) were women, 29 (58%) had a previous diagnosis of hypertension, and 35 (70%) were receiving antihypertensive therapy at hospital admission. Chronic kidney disease was present in 19 participants (38%).

**Table 1.**
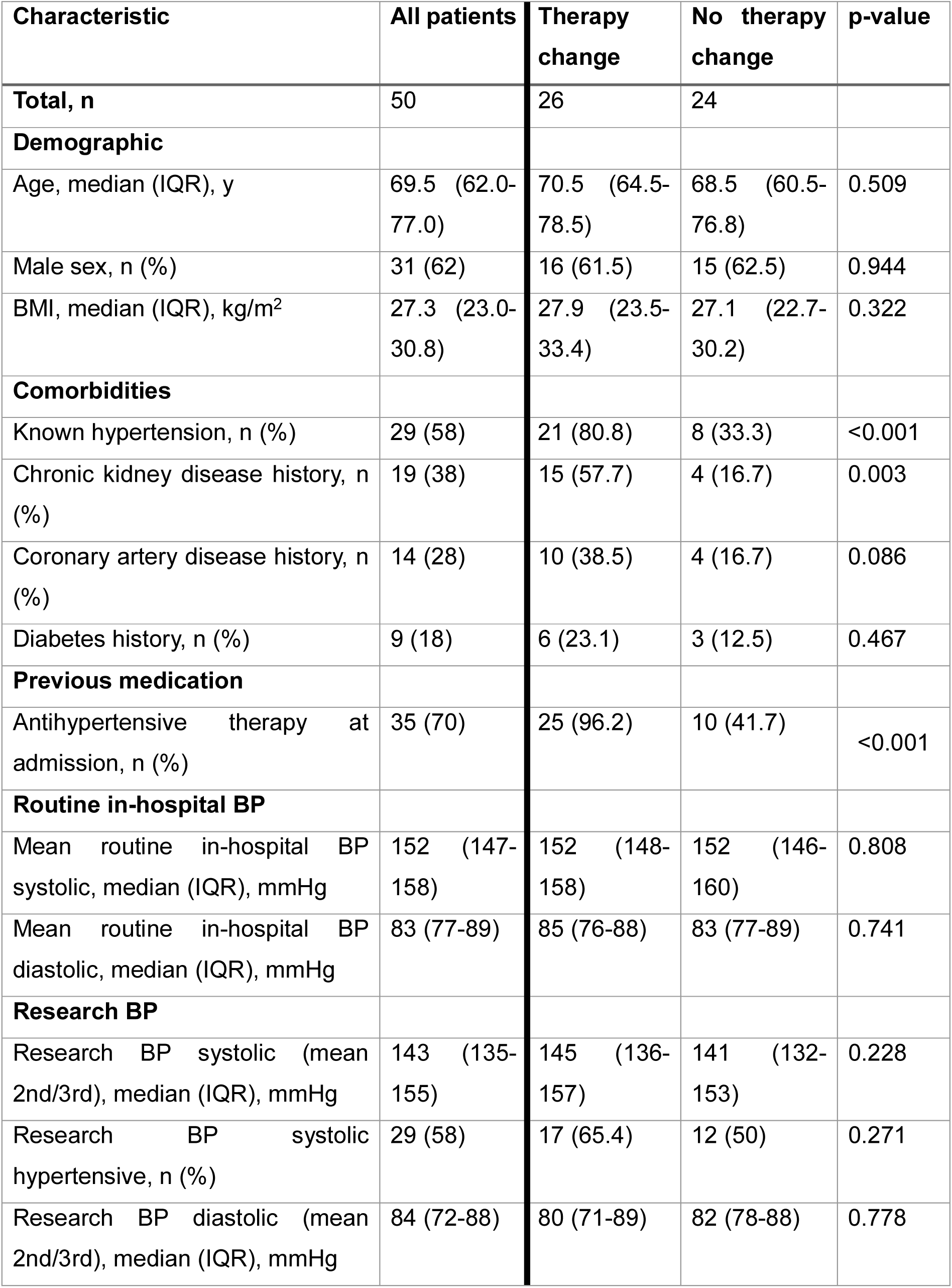
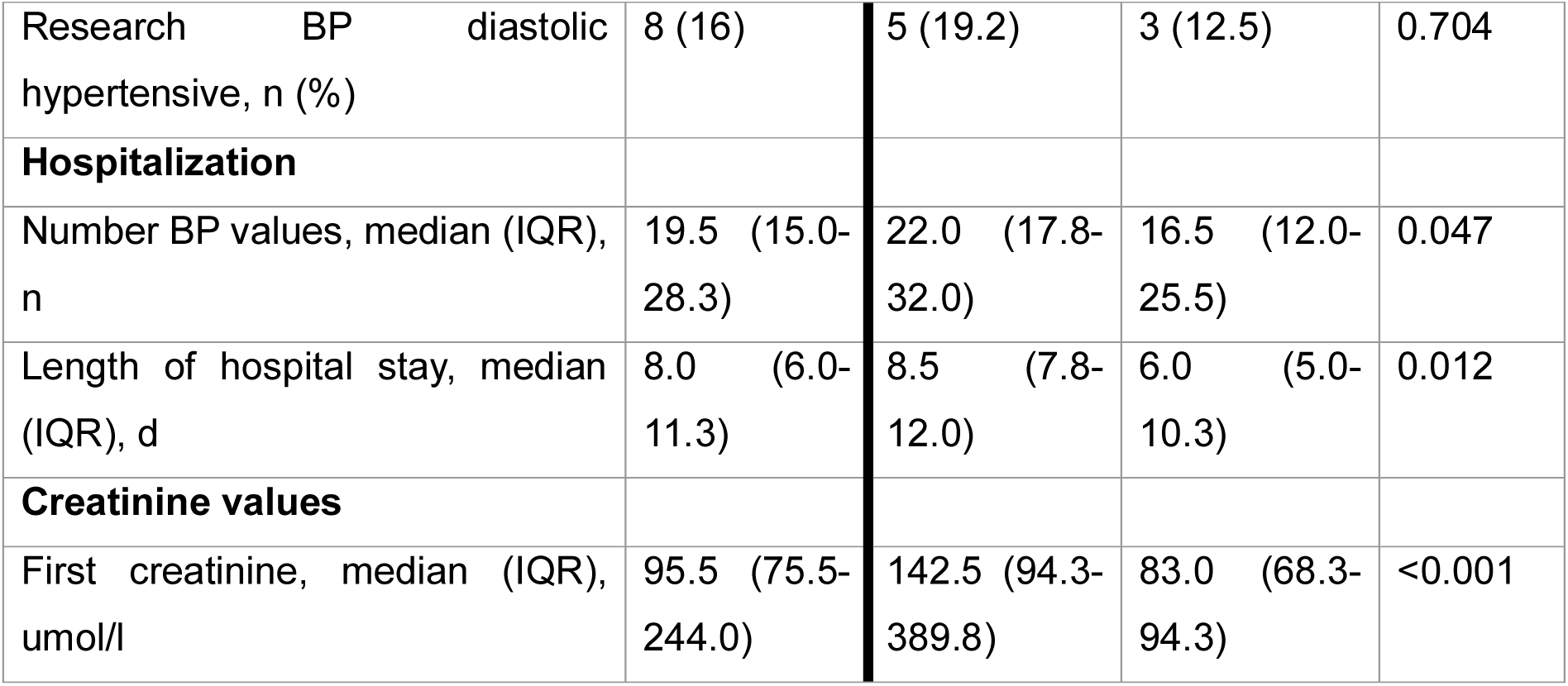
Participant characteristics according to changes in antihypertensive therapy. Data are expressed as the median (IQR = interquartile range), or n = number (%). BMI = body mass index; BP = blood pressure; d = days; y = years.

| Characteristic | All patients | Therapy change | No therapy change | p-value |
| --- | --- | --- | --- | --- |
| **Total, n** | 50 | 26 | 24 |  |
| **Demographic** |  |  |  |  |
| Age, median (IQR), y | 69.5 (62.0-77.0) | 70.5 (64.5-78.5) | 68.5 (60.5-76.8) | 0.509 |
| Male sex, n (%) | 31 (62) | 16 (61.5) | 15 (62.5) | 0.944 |
| BMI, median (IQR), kg/m<sup>2</sup> | 27.3 (23.0-30.8) | 27.9 (23.5-33.4) | 27.1 (22.7-30.2) | 0.322 |
| **Comorbidities** |  |  |  |  |
| Known hypertension, n (%) | 29 (58) | 21 (80.8) | 8 (33.3) | <0.001 |
| Chronic kidney disease history, n (%) | 19 (38) | 15 (57.7) | 4 (16.7) | 0.003 |
| Coronary artery disease history, n (%) | 14 (28) | 10 (38.5) | 4 (16.7) | 0.086 |
| Diabetes history, n (%) | 9 (18) | 6 (23.1) | 3 (12.5) | 0.467 |
| **Previous medication** |  |  |  |  |
| Antihypertensive therapy at admission, n (%) | 35 (70) | 25 (96.2) | 10 (41.7) | <0.001 |
| **Routine in-hospital BP** |  |  |  |  |
| Mean routine in-hospital BP systolic, median (IQR), mmHg | 152 (147-158) | 152 (148-158) | 152 (146-160) | 0.808 |
| Mean routine in-hospital BP diastolic, median (IQR), mmHg | 83 (77-89) | 85 (76-88) | 83 (77-89) | 0.741 |
| **Research BP** |  |  |  |  |
| Research BP systolic (mean 2nd/3rd), median (IQR), mmHg | 143 (135-155) | 145 (136-157) | 141 (132-153) | 0.228 |
| Research BP systolic hypertensive, n (%) | 29 (58) | 17 (65.4) | 12 (50) | 0.271 |
| Research BP diastolic (mean 2nd/3rd), median (IQR), mmHg | 84 (72-88) | 80 (71-89) | 82 (78-88) | 0.778 |
| Research BP diastolic hypertensive, n (%) | 8 (16) | 5 (19.2) | 3 (12.5) | 0.704 |
| **Hospitalization** |  |  |  |  |
| Number BP values, median (IQR), n | 19.5 (15.0-28.3) | 22.0 (17.8-32.0) | 16.5 (12.0-25.5) | 0.047 |
| Length of hospital stay, median (IQR), d | 8.0 (6.0-11.3) | 8.5 (7.8-12.0) | 6.0 (5.0-10.3) | 0.012 |
| **Creatinine values** |  |  |  |  |
| First creatinine, median (IQR), umol/l | 95.5 (75.5-244.0) | 142.5 (94.3-389.8) | 83.0 (68.3-94.3) | <0.001 |

### 3.2. Persistent hypertension after discharge

Four weeks after hospital discharge, 42 of 50 participants (84%; 95% CI, 70.9 – 92.8) fulfilled the predefined diagnostic ABPM criteria for hypertension in at least one measurement period (24-hour mean, awake, or asleep). Among the 21 participants without a previous diagnosis of hypertension, 18 participants (86%; 95% CI, 63.7 – 97.0; 36% of the overall cohort) had a previously unrecognized hypertension, whereas 24 of 29 participants with known hypertension (83%; 95% CI, 64.2 – 94.2, 48% of the overall cohort) had uncontrolled hypertension. Eight participants (16%) did not fulfill hypertension criteria in any ABPM measurement period.

Thirty-seven participants (74%) fulfilled hypertension criteria based on the 24-hour mean BP. Nocturnal hypertension was present in 39 of 49 participants (80%; 95% CI, 65.7 – 89.8); one participant had an insufficient number of asleep BP measurements for classification (Figure 2). Twenty-nine participants (58%) fulfilled hypertension based on systolic and/or diastolic BP across all ABPM periods; among these, 20 (40% of the overall cohort) fulfilled both systolic and diastolic hypertension criteria throughout all measurement periods, two (4%) had isolated systolic hypertension, and seven (14%) had isolated diastolic hypertension.

**Figure 2:**
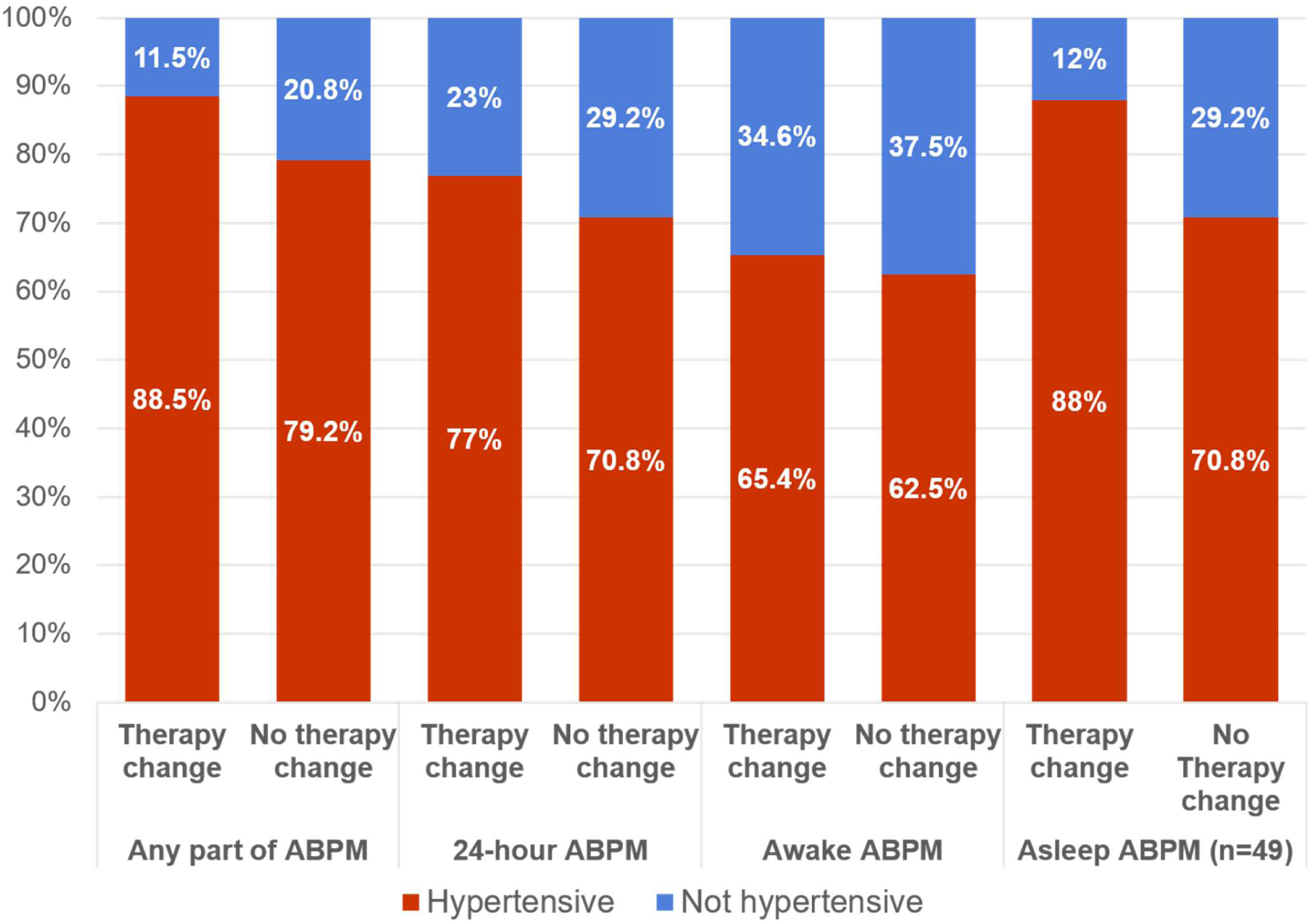
Prevalence of hypertension on follow-up ambulatory blood pressure monitoring according to changes in antihypertensive therapy during hospitalization. Bars show the proportion of participants fulfilling hypertension criteria based on 24-hour mean, awake, asleep, or any ambulatory blood pressure monitoring (ABPM) period four weeks after hospital discharge, stratified by whether antihypertensive therapy was changed during hospitalization. Percentages are calculated within each treatment group.

### 3.3. Management of elevated in-hospital BP

Antihypertensive therapy was changed during hospitalization in 26 participants (52%; 95% CI, 37.4 – 66.3). The remaining 24 participants (48%) were discharged without changes in their antihypertensive therapy. De-escalation was more common than treatment intensification: 16 participants (61.5% of those with therapy changes) underwent dose reduction or discontinuation, seven (26.9%) underwent treatment initiation or dose escalation, and three (11.5%) underwent both intensification and de-escalation during hospitalization.

Patients with and without changes in antihypertensive therapy differed descriptively in several baseline characteristics, including previous hypertension, chronic kidney disease, antihypertensive therapy at hospital admission, kidney function, and length of hospital stay (Table 1).

### 3.4. Association between in-hospital management and follow-up BP

Hypertension on follow-up ABPM was present in 23 of 26 participants (88.5%) with changes in antihypertensive therapy during hospitalization and in 19 of 24 participants (79.2%) without changes (p = 0.456; Figure 2). Hypertension prevalence according to individual ABPM periods and the distributions of systolic and diastolic ambulatory BP according to in-hospital therapy changes are shown in Figures 2 and 3, respectively. Changes in antihypertensive therapy between hospital discharge and follow-up occurred in 18 participants (36%) and are shown according to in-hospital therapy changes in Supplemental Table S2.

**Figure 3.**
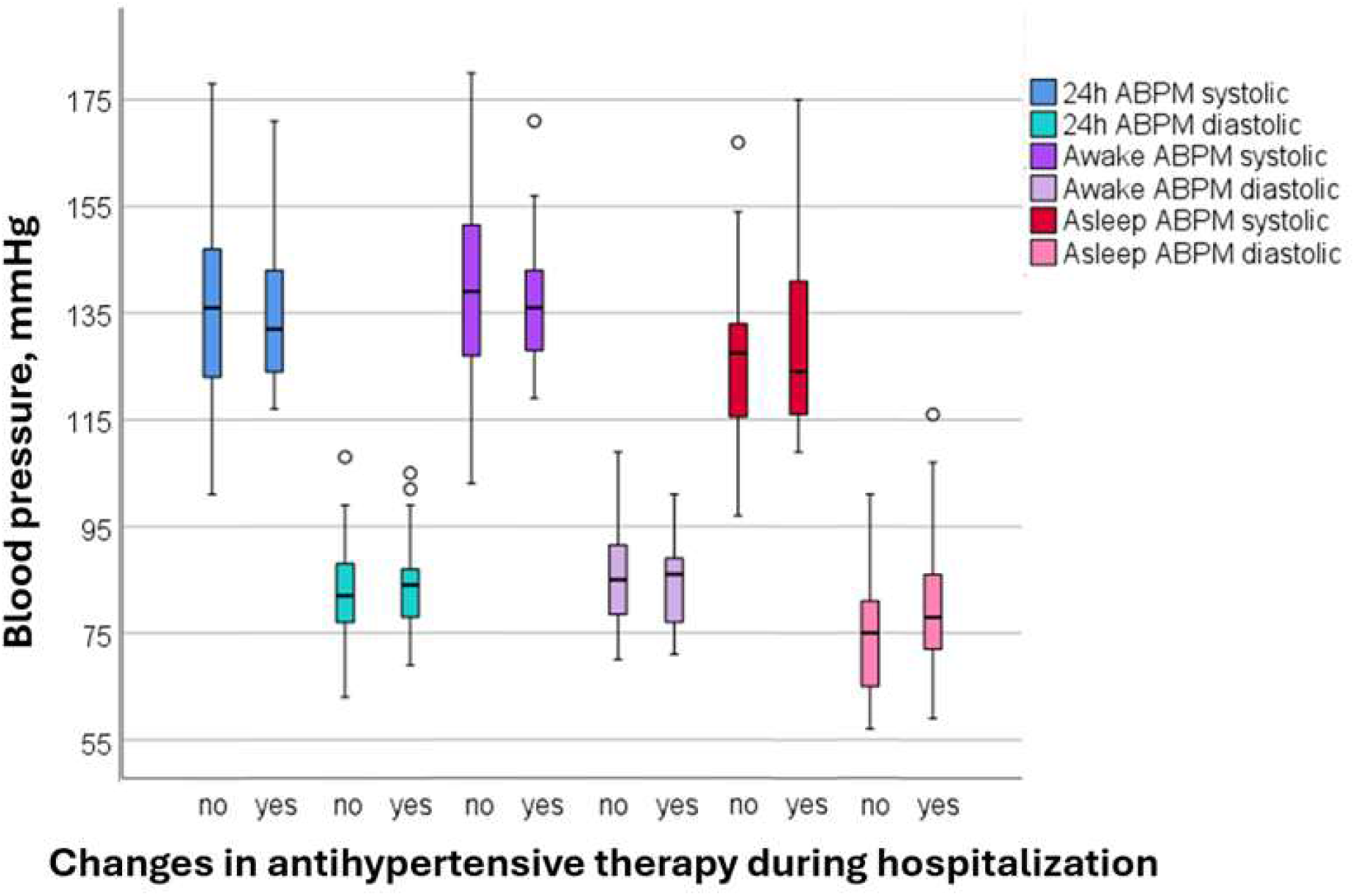
Follow-up ambulatory blood pressure measurements according to changes in antihypertensive therapy during hospitalization. Boxplots show systolic and diastolic 24-hour mean, awake, and asleep ambulatory blood pressure measurements obtained 4 weeks after hospital discharge in participants with and without changes in antihypertensive therapy during hospitalization. The horizontal line represents the median, the box the interquartile range, and the whiskers the most extreme values within 1.5 times the interquartile range; circles indicate outliers. ABPM = ambulatory blood pressure monitoring; h = hour.

### 3.5. Adverse events and rehospitalization

Seven participants (14%) experienced complications requiring rehospitalization during follow-up, including four participants with and three without changes in antihypertensive therapy during hospitalization. No myocardial infarction, intracerebral hemorrhage, ischemic stroke, syncope, or hypertensive emergency occurred. The rates of complications and rehospitalizations are shown in Table 2.

**Table 2.** Follow up and complications until 4 weeks after discharge. Data are expressed as n = number (%).

| Follow-up | All patients<br>(n = 50) | Therapy change<br>(n = 26) | No therapy<br>change (n = 24) | p-value |
| --- | --- | --- | --- | --- |
| **Complications post discharge, n (%)** | 7 (14) | 4 (15.4) | 3 (12.5) | 1.000 |
| Acute kidney injury, n (%) | 1 (2) | 1 (3.8) | 0 (0) | 1.000 |
| Rehospitalization, n (%) | 7 (14) | 4 (15.4) | 3 (12.5) | 1.000 |
| Orthostatic hypotension or presyncope, n (%) | 1 (2) | 1 (3.8) | 0 (0) | 1.000 |
| **Cause for rehospitalization** |  |  |  |  |
| Hypertensive urgency/emergency, n (%) | 0 (0) | 0 (0) | 0 (0) | 1.000 |
| Hypotension/ fall/ syncope/ presyncope, n (%) | 0 (0) | 0 (0) | 0 (0) |  |
| Cardiovascular (other), n (%) | 1 (2) | 1 (3.8) | 0 (0) |  |
| Non-cardiovascular, n (%) | 6 (12) | 3 (11.5) | 3 (12.5) |  |

### 3.6. Correlation routine in-hospital BP and standardized research BP

Routine in-hospital systolic BP was on average 10 mmHg higher than standardized research systolic BP (mean within-participant difference, 10 mmHg [95% CI, 6 - 15]; P<0.001), whereas there was no significant difference in diastolic BP (mean within-participant difference, 1 mmHg [95% CI, −1 to 4]; P=0.305; Figure 4).

**Figure 4.**
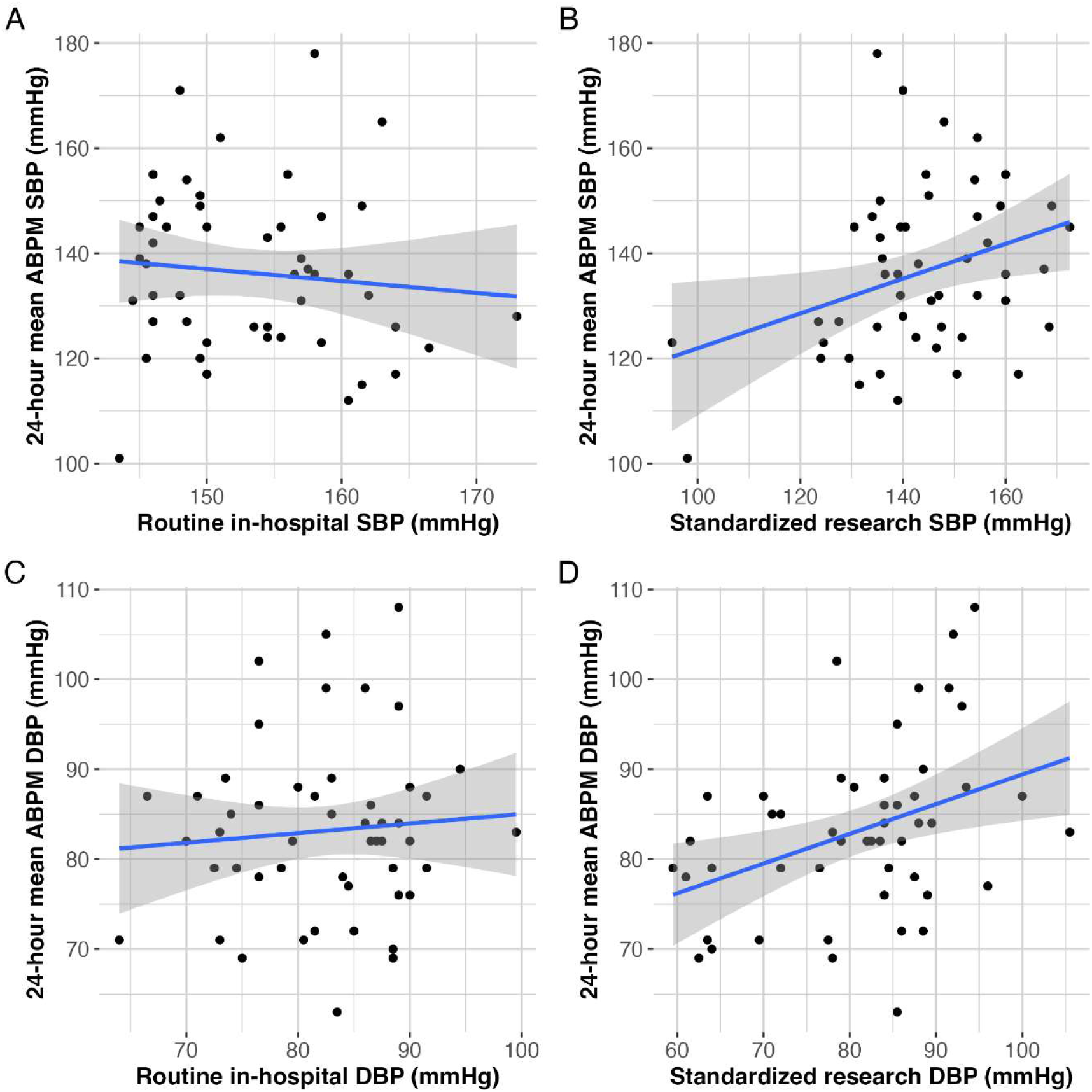
Associations of routine in-hospital and standardized research blood pressure with mean 24-hour ambulatory blood pressure. Scatterplots show the associations of routine in-hospital and standardized research BP with corresponding mean 24-hour ambulatory BP four weeks after hospital discharge. A, Routine in-hospital systolic BP. B, Standardized research systolic BP. C, Routine in-hospital diastolic BP. D, Standardized research diastolic BP. Lines indicate linear regression fits with 95% confidence intervals. ABPM = ambulatory blood pressure monitoring; SBP = systolic blood pressure, DBP = diastolic blood pressure.

Standardized research BP was modestly correlated with corresponding 24-hour mean ambulatory BP for both systolic (r=0.33 [95% CI, 0.05–0.55]; P=0.020) and diastolic BP (r=0.38 [95% CI, 0.11–0.59]; P=0.007), whereas routine in-hospital BP was not (systolic: r=−0.10 [95% CI, −0.37 to 0.18]; P=0.492; diastolic: r=0.08 [95% CI, −0.20 to 0.35]; P=0.561). Results were similar in sensitivity analyses using Spearman rank correlations.

In exploratory multivariable analyses including routine in-hospital and standardized research BP simultaneously, standardized research BP remained associated with corresponding 24-hour mean ambulatory BP for both systolic (B=0.39 [95% CI, 0.10– 0.67]; P=0.009) and diastolic BP (B=0.42 [95% CI, 0.14–0.70]; P=0.005), whereas routine in-hospital BP was not (systolic: B=−0.46 [95% CI, −1.10 to 0.18]; P=0.157; diastolic: B=−0.23 [95% CI, −0.64 to 0.18]; P=0.262).

## 4. Discussion

In this prospective INDEBP study, five principal findings emerged. First, more than four out of five participants with asymptomatic elevated routine in-hospital BP had hypertension on ABPM four weeks after discharge, indicating that elevated BP during hospitalization frequently identifies patients with persistent hypertension after discharge. Second, hypertension was highly prevalent both among participants with and without a previous diagnosis of hypertension, with previously unrecognized hypertension identified in 18 of 21 participants without a previous diagnosis and uncontrolled hypertension in 24 of 29 participants with known hypertension. Third, changes in antihypertensive therapy during hospitalization were common, with de-escalation occurring more frequently than intensification. Fourth, hypertension on follow-up ABPM was frequent irrespective of whether antihypertensive therapy had been changed during hospitalization. Finally, standardized research BP was associated with subsequent ambulatory BP, whereas routine in-hospital BP was not.

The widespread assumption that asymptomatic BP elevations during hospitalization predominantly represent transient physiological responses has influenced current clinical practice. Elevated BP is frequently attributed to pain, anxiety, sleep disturbance, or the acute illness itself, and may therefore be considered of limited clinical significance beyond the acute hospitalization^4–6, 21^. However, prospective evidence regarding the persistence of hypertension remains limited^1, 2, 7, 8^. In our cohort, more than 80% of participants were hypertensive on ABPM four weeks after discharge, when the acute hospitalization had ended. Our findings are consistent with the recently published SHINE study, which similarly demonstrated that elevated in-hospital BP can identify patients with hypertension after discharge, although SHINE focused on previously untreated individuals and the diagnostic accuracy of in-hospital BP measurements^15^. Our study extends these observations to patients both with and without known hypertension. Notably, approximately 80% of participants had nocturnal hypertension, a phenotype that may be missed by conventional office BP assessment and is associated with adverse cardiovascular and renal outcomes^22, 23^. Together, these findings support structured post-discharge BP reassessment, with ABPM providing additional information on nocturnal BP patterns.

Changes in antihypertensive therapy during hospitalization were common, but, contrary to our initial expectation, de-escalation occurred more frequently than intensification. The clinical rationale for individual treatment changes was not systematically collected and therefore cannot be determined from our data. More broadly, antihypertensive management during acute hospitalization requires consideration of BP alongside competing clinical conditions and temporary contraindications to treatment. Hypertension on follow-up ABPM remained frequent irrespective of whether antihypertensive therapy had been changed during hospitalization. This finding should not be interpreted as evidence that treatment changes are ineffective, because treatment decisions were not randomized and were subject to confounding by indication. In addition, antihypertensive therapy was frequently modified after discharge, and medication adherence was not assessed. Consequently, our findings do not establish whether or when antihypertensive therapy should be modified during hospitalization. Rather, they complement current recommendations cautioning against routine treatment changes during acute hospitalization in the absence of a hypertensive emergency by highlighting the importance of structured BP reassessment after discharge^6, 14^.

The method and timing of inpatient BP measurement may also influence the clinical information obtained from elevated in-hospital BP. Routine in-hospital systolic BP was higher than standardized research BP, and standardized research BP was associated with subsequent 24-hour ambulatory BP, whereas routine in-hospital BP was not. This finding is consistent with recent evidence demonstrating substantial variability in routine inpatient BP measurement technique and highlighting the importance of standardized measurement techniques^16, 17^. However, standardized research BP was obtained under standardized conditions and at a different time during hospitalization than routine BP measurements. Its association with follow-up ABPM may therefore reflect both the measurement protocol and its timing relative to hospital discharge, and these effects cannot be disentangled in the present study.

Several strengths of our study should be highlighted. The prospective design, predefined eligibility criteria, standardized research BP measurements, and guideline-based 24-hour ABPM after discharge allowed systematic assessment of the relationship between elevated in-hospital BP and post-discharge BP status. The inclusion of patients both with and without previously diagnosed hypertension reflects the heterogeneous population encountered in clinical practice, while prospective recording of antihypertensive treatment decisions provided insight into real-world management during hospitalization.

Several limitations should also be acknowledged. First, this analysis includes only 50 participants who had completed follow-up including ABPM, resulting in a small sample size and introducing the possibility of selection bias. Recruitment and follow-up proved challenging because many hospitalized patients were either too ill or unable to attend outpatient follow-up ABPM. Second, this was a single-center study, and patients with severe hypertension or hypertensive emergencies were excluded, limiting generalizability. Third, the study did not include a control group of hospitalized patients without elevated BP. Thus, our findings characterize the prevalence of hypertension after discharge among selected patients with elevated in-hospital BP but cannot determine how this prevalence compares with that among hospitalized patients without elevated BP. Fourth, routine in-hospital and standardized research BP measurements were obtained at different times and under different conditions; consequently, differences in their associations with subsequent ABPM cannot be attributed solely to measurement standardization. Fifth, ABPM was performed at a single follow-up visit and may therefore not fully account for biological variability in BP over time. Finally, treatment decisions were not randomized, precluding causal inference regarding changes in antihypertensive therapy, and the clinical rationale for individual treatment changes was not systematically collected. Antihypertensive therapy was also frequently modified after discharge, and medication adherence was not assessed, both of which may have influenced follow-up BP.

### 4.1. Perspectives

In this prospective study, asymptomatic elevated routine in-hospital BP identified a population with a high prevalence of hypertension on ABPM four weeks after discharge, including many patients without a previous hypertension diagnosis. These findings support using elevated in-hospital BP as a trigger for structured post-discharge BP reassessment rather than assuming that it represents a transient response to acute illness. Future studies should define the optimal timing and setting for confirming hypertension and initiating or modifying antihypertensive therapy after hospitalization.

## 5. Novelty and relevance

### 5.1. What Is New?

• More than 80% of participants with asymptomatic elevated routine in-hospital BP had previously unrecognized or uncontrolled hypertension on ABPM four weeks after discharge.

• Standardized research BP, but not routine in-hospital BP, was associated with post-discharge ambulatory BP, highlighting the potential importance of standardized BP measurement during hospitalization.

• Changes in antihypertensive therapy during hospitalization were common, with treatment de-escalation occurring more frequently than intensification.

### 5.2. What Is Relevant?

• Elevated routine in-hospital BP should not routinely be dismissed as a transient response to acute illness.

• Standardized inpatient BP measurement may help identify clinically relevant BP elevation beyond routine ward measurements.

• Elevated in-hospital BP should prompt structured post-discharge BP reassessment, ideally using ABPM.

### 5.3. Clinical/Pathophysiological Implications?

• Elevated BP during hospitalization may identify patients with hypertension persisting beyond the acute hospitalization.

• The high prevalence of nocturnal hypertension highlights the additional information provided by ABPM after discharge.

• Future studies should define the optimal timing and setting for confirming hypertension and initiating or modifying antihypertensive therapy after hospitalization.

## Data Availability

Due to ethical restrictions, the data will not be made available.

## 6. Acknowledgement

The authors thank the Department of Internal Medicine and the Medical Outpatient Department and Hypertension Clinic at University Hospital Basel, Switzerland and the study participants for their valuable contributions. All authors have reviewed and approved the final article for publication.

## Non-standard abbreviations and acronyms

ABPM: 24-hour ambulatory blood pressure monitoring
BP: blood pressure
GP: general practitioner
INDEBP: In-Hospital Detection of Elevated Blood Pressure

